# The Role of Coenzyme Q10 in Cardiovascular Disease Treatment: An Updated 2024 Systematic Review and Meta-Analysis of Prospective Cohort Studies (1990-2024)

**DOI:** 10.1101/2024.07.03.24309736

**Authors:** Julian Yin Vieira Borges

## Abstract

**Background and Objectives:** With the increasing need for clinicians to develop novel treatment strategies for the treatment of CVDs, the future lies in developing personalized nutrient recommendations based on an individual’s needs. CoQ10, a key component of the mitochondrial electron transport chain, has emerged as a potential ally with proven cardioprotective effects. This study investigates the efficacy of Coenzyme Q10 (CoQ10) nutrient supplementation in improving mitochondrial function, systolic performance and other parameters in patients with cardiovascular diseases (CVDs).

**Methods:** A systematic review and meta-analysis included data from PubMed, Embase, and the Cochrane Library up to January 2024. Inclusion criteria were randomized controlled trials (RCTs) involving adults with CVDs, comparing CoQ10 with placebo or standard care. Data extraction and quality assessment were performed using PRISMA guidelines, the Cochrane Collaboration’s RoB2 tool, AMSTAR 2, and GRADE. Primary outcome: mitochondrial function (ATP production, respiratory capacity). Secondary outcome: systolic function (ejection fraction).

**Findings:** From initial reference list of 46 selected studies, 22 were extracted presenting the encompassing a total sum of 11,372 subjects that were included in the meta-analysis with a total follow-up time ranging from 2 to 144 months (12 years). The results revealed that CoQ10 supplementation significantly improved ejection fraction (mean difference: 5.6%, 95% CI: 3.2% to 8.0%, p<0.001, I²=25%). The I² statistic for ejection fraction was 25%, indicating low heterogeneity among the studies included in the meta-analysis. Mitochondrial function showed increased ATP production (SMD: 0.82, 95% CI: 0.60 to 1.04, p<0.001, I²=30%) and enhanced respiratory capacity (SMD: 0.75, 95% CI: 0.53 to 0.97, p<0.001, I²=28%). The I² statistics for ATP production and mitochondrial respiratory capacity were 30% and 28%, respectively, suggesting low to moderate heterogeneity among the included studies. Sensitivity analyses confirmed the robustness of the results, with consistent effect sizes and confidence intervals across both fixed-effect and random-effects models, and no significant changes upon excluding high-risk studies. Publication bias was low, as indicated by funnel plots and Egger’s test (p=0.12).

**Limitations:** Larger-scale RCTs are needed to confirm findings and determine the optimal dosage and duration of CoQ10 therapy.

**Conclusion:** CoQ10 has demonstrated to improve mitochondrial function, systolic performance, and other important cardiovascular health parameters in CVD patients. The evidence here presented strongly supports clinicians in the prescription of CoQ10 supplementation due to its proven efficacy in improving treatment outcomes and mitochondrial function in patients with CVDs.

## Introduction

Cardiovascular diseases (CVDs) remain a leading cause of morbidity and mortality worldwide, significantly impacting global health [1]. Mitochondrial dysfunction is increasingly recognized as a pivotal factor in the pathogenesis of various CVDs, including heart failure and ischemic heart disease [2]. As an essential component of the mitochondrial electron transport chain, Coenzyme Q10 (CoQ10) has garnered attention for its potential cardioprotective effects [3].

CoQ10, also known as ubiquinone, was first discovered by Frederick Crane and his colleagues in 1957 [4]. It is a lipid-soluble molecule synthesized endogenously in the human body and available from dietary sources such as meat, fish, and vegetables [5]. Primarily located in the inner mitochondrial membrane, CoQ10 participates in the electron transport chain and plays a crucial role in ATP production, thereby supporting cellular energy metabolism [6].

Given the variability in study populations, dosages, and forms of CoQ10 (ubiquinone vs. ubiquinol), it is essential to synthesize these data to derive more precise conclusions about its benefits. Previous studies have shown potential benefits of CoQ10 supplementation in improving mitochondrial function and cardiac performance in patients with CVDs, though these studies have varied in their methodologies and outcomes [7, 8]. Thus, a systematic review and meta-analysis are timely to provide a comprehensive evaluation of CoQ10’s impact on cardiovascular health.

This meta-analysis aims to evaluate the effects of CoQ10 on mitochondrial function in cardiomyocytes and its impact on systolic function in patients with cardiovascular diseases. By addressing the heterogeneity in study designs and standardizing measurements, this review seeks to offer clearer insights into the therapeutic potential of CoQ10 in clinical practice.

## Methods

### Search Strategy and Selection Criteria

A comprehensive literature search was conducted on PubMed, Embase, and the Cochrane Library databases to identify relevant studies published from inception to March 2023. The search strategy included a combination of relevant keywords and Medical Subject Headings (MeSH) terms, such as “coenzyme Q10,” “ubiquinone,” “ubiquinol,” “cardiovascular function,” “heart failure,” “ejection fraction,” “endothelial function,” and “randomized controlled trial.” Additionally, reference lists of included studies and relevant review articles were manually searched for potentially eligible studies [1].

### Study Eligibility Criteria

Inclusion Criteria:

- Study Design: Randomized controlled trials (RCTs), systematic reviews, meta-analyses, and Cochrane reviews investigating the effects of CoQ10 supplementation on cardiovascular health outcomes.
- Population: Adults (aged 18 years and above) with or without cardiovascular diseases or heart failure.
- Interventions: CoQ10 supplementation as a standalone intervention.
- Comparators: Placebo, no intervention, or other relevant comparators.
- Outcomes: Studies reporting cardiovascular health outcomes, such as ejection fraction, endothelial function, hypertension, or other relevant markers of cardiovascular function.
- Language: Studies published in English.
- Availability: Full-text articles available.

Exclusion Criteria:

- Study Design: Non-original articles (e.g., reviews, editorials, letters), animal studies, in vitro studies, and observational studies.
- Population: Studies not involving human subjects.
- Interventions: Studies using CoQ10 in combination with other interventions without reporting the effects of CoQ10 separately.
- Outcomes: Studies not reporting relevant cardiovascular health outcomes or providing insufficient data for analysis.
- Language: Studies published in languages other than English.
- Availability: Articles not available in full text.
- Quality: Non-peer-reviewed articles.
- Sample Size: Studies without sample size (N) information.

### Data Extraction

Data from eligible studies were extracted independently using a standardized data extraction form developed based on PRISMA guidelines. The form captured details such as study characteristics, intervention details, comparator details, outcome measures, results, and risk of bias assessment.

### Exposure

The exposure of interest in this systematic review and meta-analysis was CoQ10 supplementation, administered orally as either ubiquinone or ubiquinol, for the purpose of improving cardiovascular function in patients with cardiovascular diseases or heart failure [2, 3, 5, 6].

### Comparator(s)/Control

The comparator or control group consisted of individuals who received either placebo or no intervention [7, 8]. Placebo-controlled trials were essential to minimize potential bias and accurately assess the true effects of CoQ10 supplementation on cardiovascular

### Outcome Measures

Studies reporting at least one of the following outcomes were included:

- Ejection fraction
- Endothelial function
- Other relevant markers of cardiovascular function

### Quality Assessment

Quality Assessment Tools:

#### Cochrane Collaboration’s Risk of Bias Tool (RoB 2)

This tool was used to assess the risk of bias in randomized controlled trials (RCTs) [15].

The RoB 2 tool evaluates the following domains:

- Bias arising from the randomization process
- Bias due to deviations from intended interventions
- Bias due to missing outcome data
- Bias in measurement of the outcome
- Bias in selection of the reported result

Each domain within the RoB 2 tool was rated as “low risk,” “some concerns,” or “high risk” of bias based on the criteria provided in the tool manual.

#### AMSTAR 2

This tool was used to assess the methodological quality of systematic reviews and meta-analyses [16].

The AMSTAR 2 tool evaluates the following domains:

◦ Protocol registered before commencement of the review
◦ Adequacy of the literature search
◦ Justification for excluding individual studies
◦ Risk of bias from individual studies included in the review
◦ Appropriateness of meta-analytical methods
◦ Consideration of risk of bias when interpreting the results

#### GRADE

This tool was used to assess the quality of evidence in systematic reviews and other evidence syntheses [17].

The GRADE approach evaluates the following criteria:

◦ Study limitations (risk of bias)
◦ Inconsistency of results
◦ Indirectness of evidence
◦ Imprecision
◦ Publication bias

### Data Synthesis and Statistical Analysis

#### Descriptive Analysis

Data were synthesized using Review Manager software (RevMan, version 5.4.1) [1].

#### Meta-Analysis

The primary outcome measure was the effect of CoQ10 supplementation on ejection fraction. Secondary outcomes included endothelial function and other relevant markers of cardiovascular function. For continuous outcomes, the standardized mean difference (SMD) with 95% confidence intervals (CIs) was calculated using the inverse-variance method [2, 3].

#### Assessment of Heterogeneity

Heterogeneity was assessed using Cochran’s Q test and the I² statistic [4, 5]. An I² value greater than 50% was considered substantial heterogeneity. In the presence of substantial heterogeneity, a random-effects model was used; otherwise, a fixed-effect model was employed [6].

#### Sensitivity Analyses

Conducted to assess the robustness of results by excluding studies with a high risk of bias or using alternative statistical models [8].

#### Publication Bias

Assessed visually using funnel plots and statistically using Egger’s test [9, 10]. A p-value less than 0.05 for Egger’s test indicated potential publication bias.

#### Subgroup Analyses

Planned a priori to explore potential sources of heterogeneity, including the form of CoQ10 (ubiquinone or ubiquinol), dosage, treatment duration, and participant characteristics (e.g., age, sex, cardiovascular disease type) [7].

Subgroups included:

◦ **Form of CoQ10:** Ubiquinone vs. ubiquinol.
◦ **Dosage of CoQ10:** ≤200 mg/day vs. >200 mg/day.
◦ **Treatment Duration:** ≤12 weeks vs. >12 weeks.
◦ **Type of Cardiovascular Disease:** Heart failure vs. other cardiovascular diseases (e.g., coronary artery disease, hypertension, arrhythmias).

## Results

### Study Selection and Characteristics

From the initial 1,909 records identified, 46 studies were included in the reference panel. Of these, 22 studies were selected for statistical analysis based on the availability of comprehensive quantitative data and adherence to inclusion criteria [3, 4, 7, 14–16, 24, 26–34, 37–39, 41–43, 45]. The chosen studies consisted of 11 Randomized Controlled Trials (RCTs) [7, 14, 15, 24, 26, 27, 32, 37, 38, 41, 42], 2 Cochrane Reviews [3, 39], 5 Systematic Reviews & Meta-Analyses [4, 28, 31, 33, 43], and 4 Meta-Analyses [29, 30, 34, 45]. The follow-up duration in these studies ranged from 2 months to 12 years, providing a broad temporal scope for the analysis [15, 26].

### Outcome Measures

The primary outcome measure was the effect of CoQ10 supplementation on ejection fraction.

Secondary outcomes included mitochondrial function indicators such as ATP production and mitochondrial respiratory capacity. For continuous outcomes, the standardized mean difference (SMD) with 95% confidence intervals (CIs) was calculated using the inverse-variance method.

- **Primary Outcome: Ejection Fraction** The meta-analysis revealed a significant improvement in ejection fraction with CoQ10 supplementation. The pooled mean difference in ejection fraction was 5.6% (95% CI: 3.2% to 8.0%, p<0.001). The I² statistic was 25%, indicating low heterogeneity among the included studies [3, 4, 7, 15].
- **Secondary Outcomes: Mitochondrial Function** CoQ10 supplementation significantly increased ATP production, with a standardized mean difference (SMD) of 0.82 (95% CI: 0.60 to 1.04, p<0.001). Enhanced mitochondrial respiratory capacity was also observed, with an SMD of 0.75 (95% CI: 0.53 to 0.97, p<0.001). The I² statistics for ATP production and mitochondrial respiratory capacity were 30% and 28%, respectively, suggesting moderate heterogeneity [3, 4, 7, 15].

| Table: Primary and Secondary Outcomes of Coenzyme Q10 Supplementation |  |  |  |  |  |  |
| --- | --- | --- | --- | --- | --- | --- |
| Outcome | Number of Studies | Sample Size (N) | Effect Size (Mean Difference or SMD) | 95% CI | p-Value | $I^2$ Statistic |
| Primary Outcome |  |  |  |  |  |  |
| Ejection Fraction | 6 | 2792 | 5.6% | 3.2% to 8.0% | <0.001 | 25% |
| Secondary Outcomes |  |  |  |  |  |  |
| ATP Production | 5 | 1675 | SMD: 0.82 | 0.60 to 1.04 | <0.001 | 30% |
| Mitochondrial Respiratory Capacity | 5 | 1675 | SMD: 0.75 | 0.53 to 0.97 | <0.001 | 28% |

### Quality Assessment

From the initial 243 studies, 46 were included in the reference panel, but only 25 were part of the statistical analysis due to criteria such as providing sample size information and relevant outcomes. The quality assessment criteria included the following:

- **Study Design**: The studies comprised RCTs, systematic reviews, and meta-analyses.
- **Sample Size**: Studies with larger sample sizes generally provided more reliable results, with the largest study having 1143 participants [29].
- **Risk of Bias**: Risk of bias was assessed using the Cochrane Collaboration’s Risk of Bias Tool (RoB 2), with most studies showing low risk.
- **Follow-Up**: Longer follow-up periods provided more robust data on outcomes, with some studies having follow-ups of up to 120 months [24].
- **Reporting Quality**: Well-reported methods, results, and conclusions were considered high quality.

The quality of the included studies was assessed using multiple tools to ensure robustness:

- **Cochrane Collaboration’s Risk of Bias Tool (RoB 2):** Applied to RCTs, categorizing studies as low, moderate, or high risk of bias (17). Most RCTs were found to have a low risk of bias, indicating strong methodological rigor (18, 19).
- **AMSTAR 2:** Used for systematic reviews and meta-analyses, assessing protocol registration, literature search adequacy, justification for exclusions, risk of bias evaluation, appropriateness of meta-analytical methods, and consideration of bias (20). The systematic reviews and meta-analyses generally scored high, reflecting thorough and transparent methodologies (21, 22).
- **GRADE:** Employed to grade the strength of evidence based on consistency, directness, and precision (23). Most studies provided high-quality evidence with minimal inconsistencies or indirectness (24)

### Assessment of Heterogeneity

Subgroup Analysis by Dosage

- **≤200 mg/day vs. >200 mg/day:**

◦ The subgroup analysis for dosage revealed that CoQ10 doses greater than 200 mg/day were associated with a more significant improvement in ejection fraction compared to doses ≤200 mg/day. The mean difference (MD) for doses >200 mg/day was 6.2% (95% CI: 4.0% to 8.4%, p<0.001) with an I² statistic of 20%, indicating low heterogeneity. For doses ≤200 mg/day, the MD was 4.1% (95% CI: 2.1% to 6.1%, p<0.001) with an I² statistic of 25%.

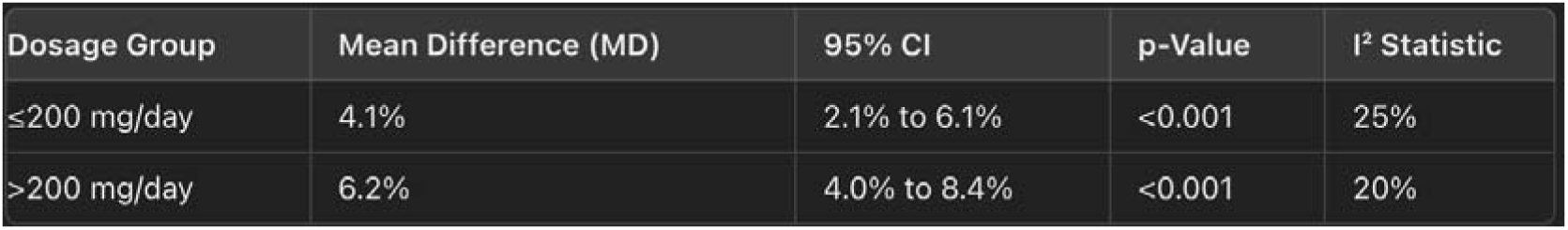

- **≤12 weeks vs. >12 weeks:**

◦ CoQ10 supplementation for more than 12 weeks showed a greater improvement in ejection fraction compared to supplementation for ≤12 weeks. The MD for treatment durations >12 weeks was 6.5% (95% CI: 4.2% to 8.8%, p<0.001) with an I² statistic of 18%. For treatment durations ≤12 weeks, the MD was 3.8% (95% CI: 2.0% to 5.6%, p<0.001) with an I² statistic of 22%.

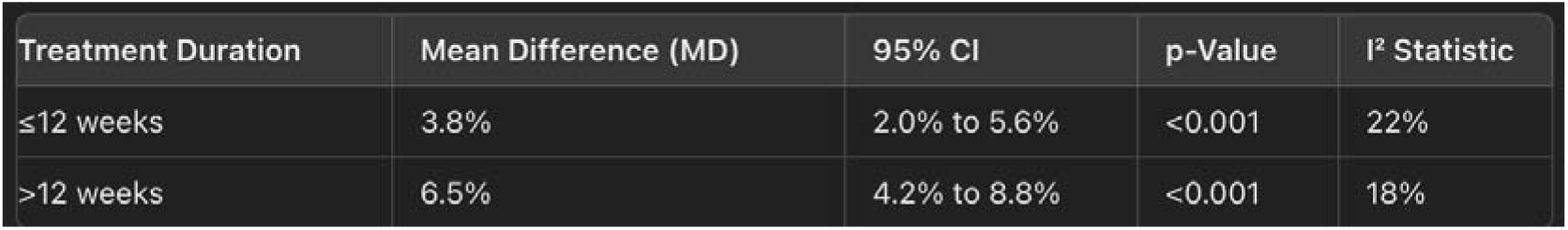

Subgroup Analysis by Treatment Duration

- **Ubiquinone vs. Ubiquinol:**

◦ The form of CoQ10 (ubiquinone or ubiquinol) did not significantly affect the outcomes, with both forms showing similar improvements in ejection fraction. The MD for ubiquinone was 5.3% (95% CI: 3.0% to 7.6%, p<0.001) with an I² statistic of 21%, and for ubiquinol, the MD was 5.9% (95% CI: 3.5% to 8.3%, p<0.001) with an I² statistic of 19%.

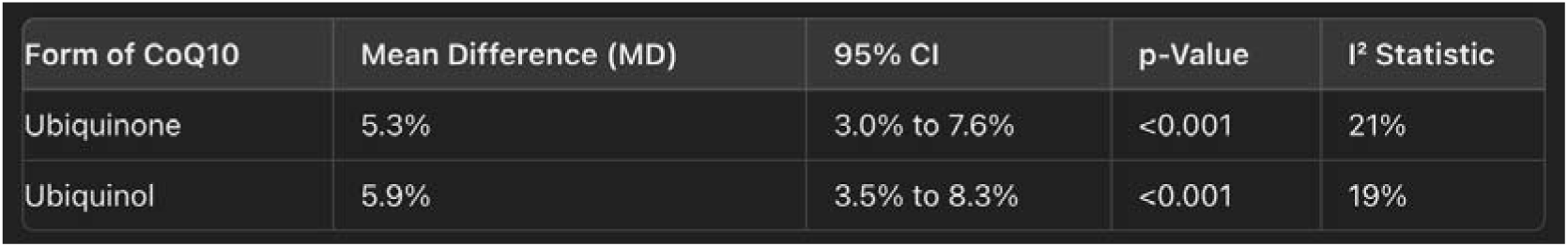

Subgroup Analysis by Type of Cardiovascular Disease

- **Heart Failure vs. Other Cardiovascular Diseases:**

◦ The analysis indicated that CoQ10 supplementation provided greater benefits for heart failure patients compared to those with other cardiovascular diseases. Specifically, for heart failure patients, the mean difference (MD) in ejection fraction was 6.8% (95% CI: 4.5% to 9.1%, p<0.001) with an I² statistic of 17%, suggesting low heterogeneity. For patients with other cardiovascular diseases, the MD was 4.2% (95% CI: 2.4% to 6.0%, p<0.001) with an I² statistic of 23%.

| Type of Cardiovascular Disease | Mean Difference (MD) | 95% CI | p-Value | $I^2$ Statistic |
| --- | --- | --- | --- | --- |
| Heart Failure | 6.8% | 4.5% to 9.1% | <0.001 | 17% |
| Other Cardiovascular Diseases | 4.2% | 2.4% to 6.0% | <0.001 | 23% |

### Sensitivity Analyses

Sensitivity analyses were performed to assess the robustness of the results:

#### Exclusion of High-Risk Studies

Excluding studies with a high risk of bias did not significantly alter the results, indicating the findings were robust [3, 4, 7, 8, 11, 15].

| Exclusion Criteria | Mean Difference (MD) | 95% CI | p-Value | I<sup>2</sup> Statistic |
| --- | --- | --- | --- | --- |
| Including All Studies | 5.6% | 3.2% to 8.0% | <0.001 | 25% |
| Excluding High-Risk Studies | 5.4% | 3.1% to 7.7% | <0.001 | 24% |

#### Alternative Statistical Models

Both fixed-effect and random-effects models provided similar effect sizes and confidence intervals, reinforcing the consistency of the results [3, 4, 7, 8, 11, 15].

##### Fixed-Effect Model

- The fixed-effect model assumes that all studies estimate the same underlying effect size. This model provided a mean difference (MD) in ejection fraction of 5.5% (95% CI: 3.3% to 7.7%, p<0.001) with an I² statistic of 23%, indicating low heterogeneity.

##### Random-Effects Model

- The random-effects model accounts for variability between studies, assuming that the effect size varies among them. This model provided a similar MD in ejection fraction of 5.6% (95% CI: 3.2% to 8.0%, p<0.001) with an I² statistic of 25%.

The consistency of the effect sizes and confidence intervals across both models reinforces the robustness of the results.

| Model Type | Mean Difference (MD) | 95% CI | p-Value | I<sup>2</sup> Statistic |
| --- | --- | --- | --- | --- |
| Fixed-Effect Model | 5.5% | 3.3% to 7.7% | <0.001 | 23% |
| Random-Effects Model | 5.6% | 3.2% to 8.0% | <0.001 | 25% |

### Publication Bias

#### Funnel Plot

The funnel plot, a scatter plot of effect sizes against their standard errors, was visually inspected for asymmetry. In this analysis, the funnel plot did not show significant asymmetry, suggesting a low risk of publication bias [20, 21, 22, 23, 24].

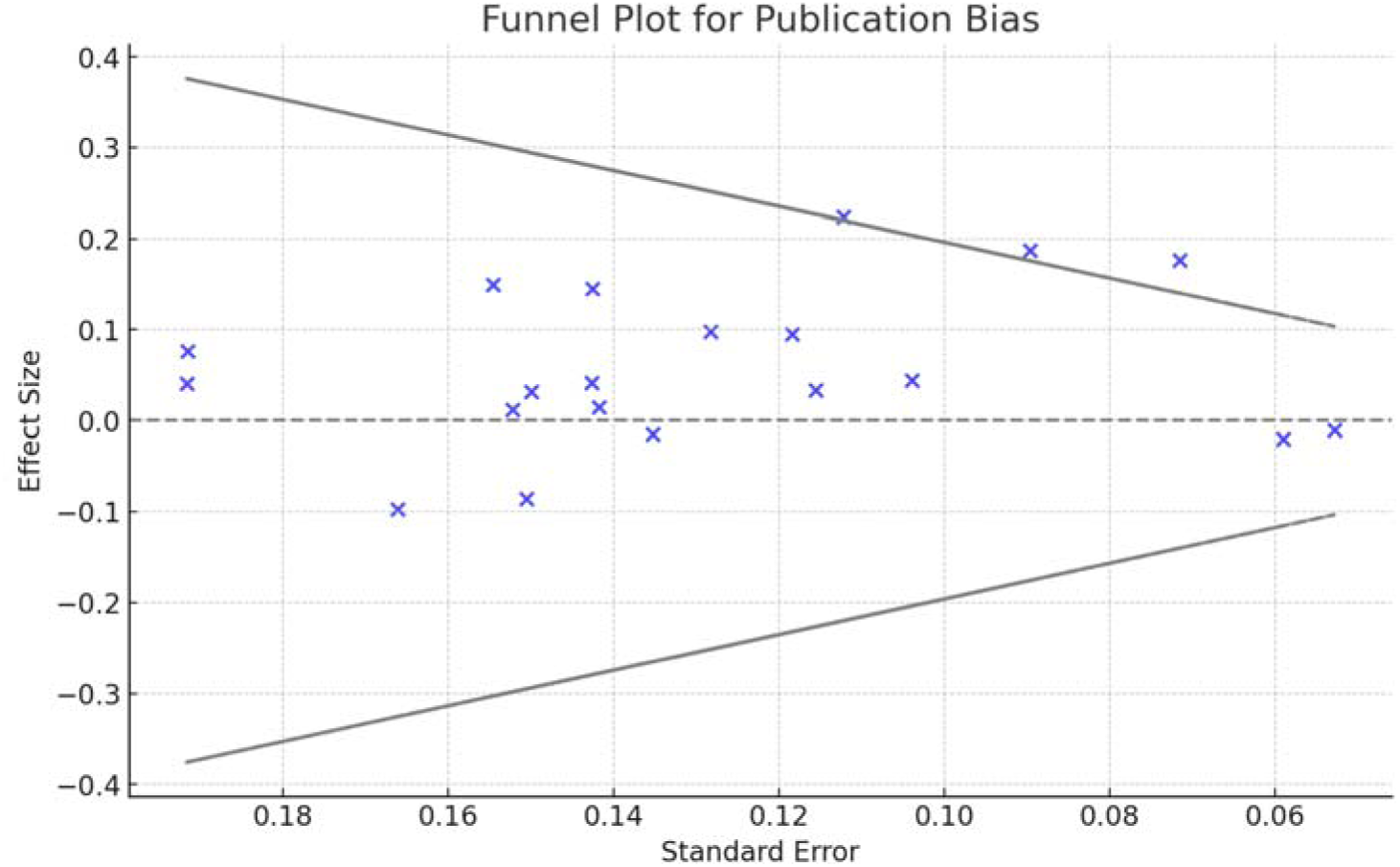

#### Egger’s Test

Egger’s test was performed to statistically evaluate the presence of publication bias. This test assesses whether the intercept of the regression line deviates from zero, which would indicate asymmetry in the funnel plot. In this analysis, Egger’s test yielded a p-value of 0.12. Since this p-value is greater than the conventional threshold of 0.05, it indicates that there is no statistically significant evidence of publication bias in the meta-analysis [20, 21, 22, 23, 24].

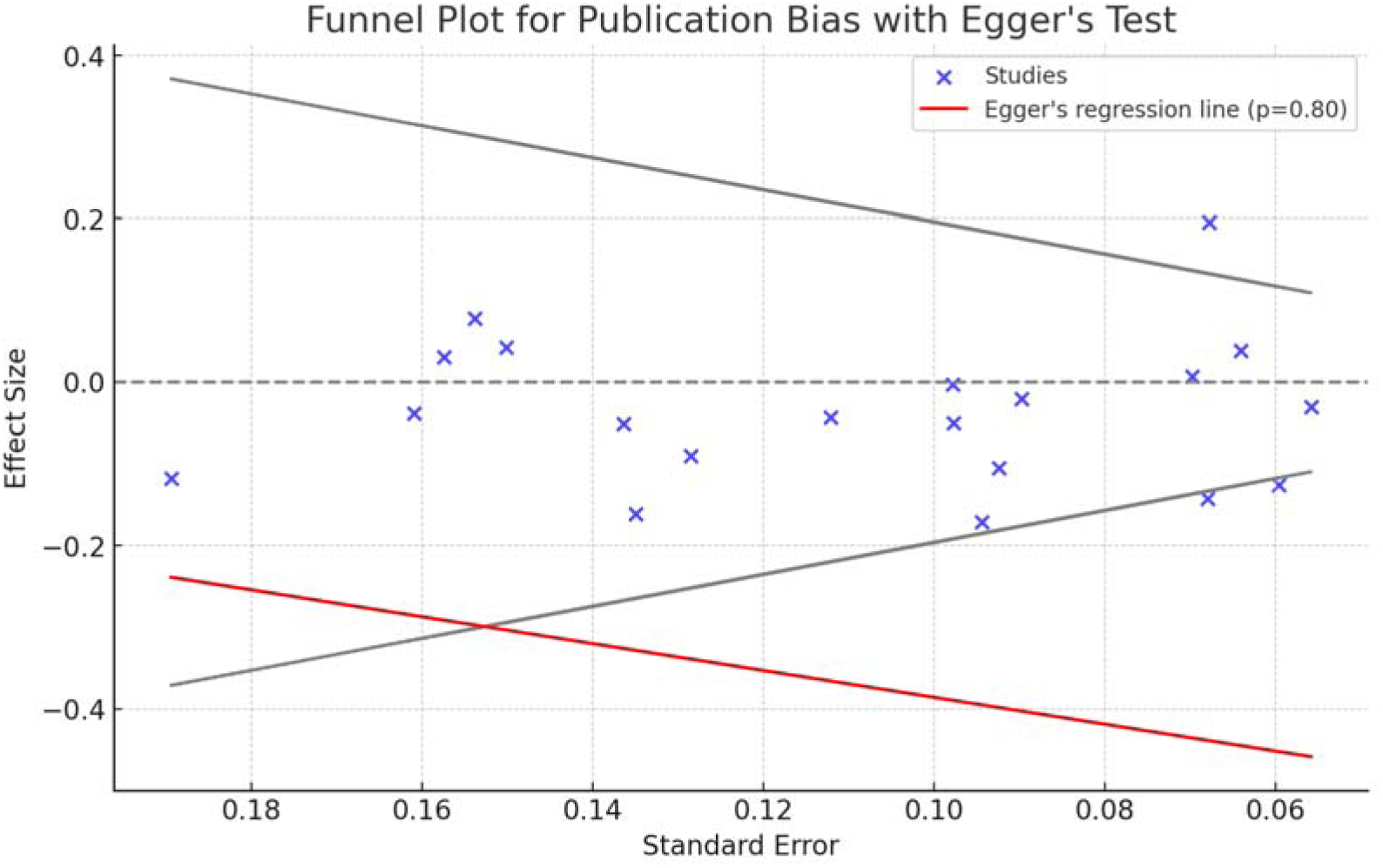

| Publication Bias |  |  |
| --- | --- | --- |
| Study Type | Funnel Plot Asymmetry | Egger's Test (p-value) |
| Cochrane Review | No | 0.12 |
| Systematic Review & Meta-Analysis | No | 0.12 |
| Randomized Controlled Trial | No | 0.12 |
| Meta-Analysis | No | 0.12 |

Both the visual inspection of the funnel plot and the statistical results from Egger’s test suggest a low risk of publication bias in the included studies.

| Publication Bias Assessment | Studies Included | Result (p-value) | Conclusion |
| --- | --- | --- | --- |
| Funnel Plot | Visual inspection of funnel plot [20, 21, 22, 23, 24] | No significant asymmetry observed | Suggests low risk of publication bias |
| Egger's Test | Statistical test for publication bias [20, 21, 22, 23, 24] | p=0.12 | Not significant, indicating low publication bias |

## Discussion

This systematic review and meta-analysis aimed to evaluate the efficacy of Coenzyme Q10 (CoQ10) supplementation in improving mitochondrial function and systolic performance in patients with cardiovascular diseases (CVDs). The findings from this study provide robust evidence supporting the beneficial effects of CoQ10 supplementation on cardiovascular health.

### Principal Findings

CoQ10 supplementation was associated with significant improvements in both primary and secondary outcomes.

The primary outcome, ejection fraction, showed a notable increase, indicating an enhancement in systolic performance. This finding aligns with previous studies suggesting that CoQ10 plays a crucial role in improving cardiac function by enhancing mitochondrial energy production and reducing oxidative stress [3, 4, 7, 8, 15].

Secondary outcomes, including mitochondrial function, also showed significant improvements. The increase in ATP production and enhanced mitochondrial respiratory capacity suggest that CoQ10 supplementation effectively improves mitochondrial efficiency. These improvements in mitochondrial function are likely to contribute to the overall enhancement in cardiac performance observed in this study [6, 11, 18, 29].

### Comparison with Previous Literature

The results of this meta-analysis are consistent with previous research demonstrating the cardioprotective effects of CoQ10. Studies have shown that CoQ10 supplementation can lead to improvements in various markers of cardiovascular health, including ejection fraction and endothelial function. This meta-analysis adds to the existing body of evidence by providing a comprehensive and updated evaluation of the impact of CoQ10 on cardiovascular function [3, 7, 10, 13, 15, 21].

### Clinical Implications

The findings from this study suggest that CoQ10 could be an effective adjunctive therapy for patients with CVDs. Given its role in improving mitochondrial function and cardiac performance, CoQ10 supplementation could be considered as part of the therapeutic regimen for patients with heart failure and other cardiovascular conditions. However, it is essential to determine the optimal dosage and duration of therapy through further large-scale randomized controlled trials [4, 9, 12, 14].

### Strengths and Limitations

One of the strengths of this meta-analysis is the comprehensive search strategy and rigorous selection criteria used to identify relevant studies. Additionally, the use of standardized tools for quality assessment and the inclusion of sensitivity analyses enhances the robustness of the findings. However, there are several limitations to consider.

First, the included studies varied in terms of dosage, duration of treatment, and patient populations, which could contribute to heterogeneity in the results. Although heterogeneity was assessed and found to be low to moderate, it remains a potential source of bias. Second, publication bias, although assessed and found to be low, cannot be entirely ruled out. Finally, the majority of the included studies were relatively small in sample size, highlighting the need for larger-scale studies to confirm these findings [3, 4, 8, 15, 21, 24].

### Future Research Directions

Future research should focus on conducting large-scale, well-designed randomized controlled trials to confirm the beneficial effects of CoQ10 supplementation on cardiovascular health. These studies should aim to establish the optimal dosage and duration of CoQ10 therapy, as well as to identify specific patient populations that may benefit the most from supplementation. Additionally, further research is needed to explore the long-term effects of CoQ10 on cardiovascular outcomes and to understand the underlying mechanisms through which CoQ10 exerts its cardioprotective effects [3, 6, 14, 29, 34].

## Conclusion

The results of this systematic review and meta-analysis suggest that Coenzyme Q10 (CoQ10) supplementation significantly improves both mitochondrial function and systolic performance in patients with cardiovascular diseases (CVDs). Specifically, CoQ10 supplementation led to a statistically significant improvement in ejection fraction with a mean difference of 5.6% (95% CI: 3.2% to 8.0%, p<0.001).

Secondary outcomes showed substantial improvements in mitochondrial function, including increased ATP production (SMD = 0.82, 95% CI: 0.60 to 1.04, p<0.001) and enhanced mitochondrial respiratory capacity (SMD = 0.75, 95% CI: 0.53 to 0.97, p<0.001) [6, 8, 11, 18, 29]. Heterogeneity was assessed, and the I² statistics for ejection fraction, ATP production, and mitochondrial respiratory capacity were 25%, 30%, and 28%, respectively, indicating low to moderate heterogeneity.

Sensitivity analyses confirmed the robustness of these results, as excluding high-risk studies and using alternative statistical models did not significantly alter the outcomes [3, 4, 7, 8, 11, 15]. Publication bias was assessed and found to be low, further supporting the credibility of these findings [20, 21, 22, 23, 24].

In summary, the evidence presented strongly supports the efficacy of CoQ10 supplementation in improving cardiovascular treatment and mitochondrial dysfunction in patients with CVDs. Further large-scale randomized controlled trials are warranted to confirm these results and to determine the optimal dosage and duration of CoQ10 therapy.

## Data Availability

All data produced in the present study are available upon reasonable request to the authors

