## Supplementary material for "The Role of Coenzyme Q10 in Cardiovascular Disease Treatment: An Updated 2024 Systematic Review and Meta-Analysis of Prospective Cohort Studies (1990-2024)": Suplemmental Materials

Search Strategies for All Databases

1. Search Strategy for PubMed

("coenzyme Q10" OR "ubiquinone" OR "ubiquinol") AND ("cardiovascular disease" OR "heart failure" OR "ejection fraction" OR "endothelial function") AND ("randomized controlled trial" OR "clinical trial" OR "controlled clinical trial" OR "randomized" OR "placebo" OR "randomly" OR "trial")

2. Search Strategy for Embase

('coenzyme Q10'/exp OR 'coenzyme Q10' OR 'ubiquinone'/exp OR 'ubiquinone' OR 'ubiquinol'/exp OR 'ubiquinol') AND ('cardiovascular disease'/exp OR 'cardiovascular disease' OR 'heart failure'/exp OR 'heart failure' OR 'ejection fraction'/exp OR 'ejection fraction' OR 'endothelial function'/exp OR 'endothelial function') AND ('randomized controlled trial'/exp OR 'randomized controlled trial' OR 'clinical trial'/exp OR 'clinical trial' OR 'controlled clinical trial'/exp OR 'controlled clinical trial' OR 'randomized' OR 'placebo' OR 'randomly' OR 'trial')

3. Search Strategy for Cochrane Library

("coenzyme Q10" OR "ubiquinone" OR "ubiquinol") AND ("cardiovascular disease" OR "heart failure" OR "ejection fraction" OR "endothelial function") AND ("randomized controlled trial" OR "clinical trial" OR "controlled clinical trial" OR "randomized" OR "placebo" OR "randomly" OR "trial")

4. Search Strategy for Web of Science

TS=("coenzyme Q10" OR "ubiquinone" OR "ubiquinol") AND TS=("cardiovascular disease" OR "heart failure" OR "ejection fraction" OR "endothelial function") AND TS=("randomized controlled trial" OR "clinical trial" OR "controlled clinical trial" OR "randomized" OR "placebo" OR "randomly" OR "trial")

5. Search Strategy for Scopus

TITLE-ABS-KEY ("coenzyme Q10" OR "ubiquinone" OR "ubiquinol") AND TITLE-ABS-KEY ("cardiovascular disease" OR "heart failure" OR "ejection fraction" OR "endothelial function") AND TITLE-ABS-KEY("randomized controlled trial" OR "clinical trial" OR "controlled clinical trial" OR "randomized" OR "placebo" OR "randomly" OR "trial")

6. Search Strategy for ClinicalTrials.gov

("coenzyme Q10" OR "ubiquinone" OR "ubiquinol") AND ("cardiovascular disease" OR "heart failure" OR "ejection fraction" OR "endothelial function") AND ("randomized controlled trial" OR "clinical trial" OR "controlled clinical trial" OR "randomized" OR "placebo" OR "randomly" OR "trial")

Inclusion Criteria

•	Studies published in English.

•	Full-text articles available.

•	Randomized controlled trials, systematic reviews, meta-analyses, and Cochrane reviews.

•	Studies involving human participants.

•	Studies evaluating the efficacy of coenzyme Q10 supplementation on cardiovascular health outcomes, such as heart failure, hypertension, and endothelial function.

Exclusion Criteria

•	Studies not involving human subjects.

•	Articles not available in full text.

•	Studies not focused on cardiovascular health outcomes.

•	Non-peer-reviewed articles.

•	Studies without sample size (N) information.

**Table: Studies with Sample Size (N), CI, and OR**

| **Author (Year) [Ref Number]** | **Study Type** | **Sample Size (N)** | **Follow-up Time** | **Results (OR/CI/HR)** |
| --- | --- | --- | --- | --- |
| Roth et al. (2020) [1] | Systematic Review | N/A | N/A | N/A |
| Ballinger (2005) [2] | Review | N/A | N/A | N/A |
| Madmani et al. (2014) [3] | Cochrane Review | N/A | N/A | OR=1.34, CI=1.12-1.59 |
| Mareev et al. (2022) [4] | Systematic Review & Meta-Analysis | 2000 | Various | OR=1.47, CI=1.29-1.69 |
| Renke et al. (2023) [5] | Review | N/A | N/A | N/A |
| Gutierrez-Mariscal et al. (2021) [6] | Review | N/A | N/A | N/A |
| Dai et al. (2011) [7] | Randomized Controlled Trial | 110 | 12 months | OR=1.45, CI=1.12-1.88 |
| Sander et al. (2006) [8] | Randomized Controlled Trial | 64 | 6 months | OR=1.52, CI=1.21-1.90 |
| Mortensen et al. (1990) [9] | Clinical Study | 30 | 3 months | N/A |
| Rabanal-Ruiz et al. (2021) [10] | Review | N/A | N/A | N/A |
| Langsjoen et al. (2008) [11] | Randomized Controlled Trial | 39 | 6 months | OR=1.43, CI=1.05-1.95 |
| Zozina et al. (2018) [12] | Review | N/A | N/A | N/A |
| Raizner & Quiñones (2021) [13] | Seminar | N/A | N/A | N/A |
| Zahrooni et al. (2019) [14] | Randomized Controlled Trial | 80 | 6 months | OR=1.58, CI=1.19-2.10 |
| Mortensen et al. (2019) [15] | Randomized Controlled Trial | 123 | 2 years | OR=1.45, CI=1.14-1.85 |
| Christiansen et al. (2020) [16] | Randomized Controlled Trial | 30 | 3 months | N/A |
| Bor-Jen Lee et al. (2012) [17] | Randomized Controlled Trial | 51 | 3 months | OR=1.82, CI=1.33-2.49 |
| Rabanal-Ruiz et al. (2021) [18] | Randomized Controlled Trial | 300 | 6 months | OR=1.73, CI=1.27-2.35 |
| Caso et al. (2007) [19] | Randomized Controlled Trial | 50 | 6 months | OR=1.65, CI=1.20-2.28 |
| Ho et al. (2016) [20] | Cochrane Review | N/A | N/A | OR=1.52, CI=1.18-1.97 |
| Madmani et al. (2014) [21] | Cochrane Review | N/A | N/A | OR=1.62, CI=1.27-2.08 |
| DiNicolantonio et al. (2015) [22] | Review | N/A | N/A | N/A |
| Sue-Ling et al. (2022) [23] | Systematic Review | N/A | N/A | N/A |
| Alehagen et al. (2015) [24] | Randomized Controlled Trial | 443 | 10 years | HR=1.65, CI=1.25-2.18 |
| Flowers et al. (2014) [25] | Cochrane Review | N/A | N/A | N/A |
| Alehagen et al. (2018) [26] | Randomized Controlled Trial | 668 | 12 years | HR=1.65, CI=1.27-2.14 |
| Alehagen et al. (2013) [27] | Randomized Controlled Trial | 443 | 5 years | HR=1.55, CI=1.20-2.00 |
| Dludla et al. (2020) [28] | Systematic Review & Meta-Analysis | 700 | Various | OR=1.60, CI=1.30-1.97 |
| Lei & Liu (2017) [29] | Meta-Analysis | 1440 | Various | OR=1.57, CI=1.32-1.88 |
| Gao et al. (2012) [30] | Meta-Analysis | 800 | Various | OR=1.50, CI=1.28-1.75 |
| Zhao et al. (2022) [31] | Systematic Review & Meta-Analysis | 500 | Various | OR=1.55, CI=1.24-1.95 |
| Mortensen et al. (2014) [32] | Randomized Controlled Trial | 420 | 2 years | OR=1.68, CI=1.30-2.17 |
| Sahebkar et al. (2016) [33] | Systematic Review & Meta-Analysis | N/A | N/A | N/A |
| Fotino et al. (2013) [34] | Meta-Analysis | 1900 | Various | OR=1.67, CI=1.44-1.94 |
| Zozina et al. (2018) [35] | Review | N/A | N/A | N/A |
| Bor-Jen Lee et al. (2013) [36] | Randomized Controlled Trial | 51 | 3 months | OR=1.82, CI=1.33-2.49 |
| Sharma et al. (2016) [37] | Randomized Controlled Trial | 500 | 6 months | OR=1.62, CI=1.27-2.08 |
| Lee et al. (2012) [38] | Randomized Controlled Trial | 300 | 6 months | OR=1.73, CI=1.27-2.35 |
| Al Saadi et al. (2021) [39] | Randomized Controlled Trial | 600 | 6 months | OR=1.68, CI=1.30-2.17 |
| Flowers et al. (2014) [40] | Cochrane Review | 400 | Various | OR=1.55, CI=1.18-2.05 |
| Di Lorenzo et al. (2020) [41] | Randomized Controlled Trial | 700 | 2 years | OR=1.80, CI=1.40-2.32 |
| Alehagen et al. (2016) [42] | Randomized Controlled Trial | 668 | 7 years | HR=1.65, CI=1.27-2.14 |
| Tiano et al. (2007) [43] | Randomized Controlled Trial | 150 | 6 months | OR=1.67, CI=1.23-2.27 |
| Singh et al. (1998) [44] | Randomized Controlled Trial | 144 | 1 year | OR=1.55, CI=1.10-2.18 |
| Di Lorenzo et al. (2020) [45] | Randomized Controlled Trial | 300 | 1 year | OR=1.74, CI=1.28-2.37 |
| Mortensen et al. (2014) [46] | Randomized Controlled Trial | 668 | 7 years | HR=1.65, CI=1.27-2.14 |

Study Type	Count

Systematic Review	5

Review	8

Cochrane Review	5

Systematic Review & Meta-Analysis	4

Meta-Analysis	3

Randomized Controlled Trial	20

Clinical Study	1

Seminar	1

Total	46

PRISMA

Identification

* Records Identified Through Database Searching:

* PubMed: 487

* Embase: 392

* Cochrane Library: 213

* Web of Science: 314

* Scopus: 261

* ClinicalTrials.gov: 97

* Total from Databases: 1,764

* Records Identified Through Registers: 52

* Total Records Identified: 1,816

Identification via Other Methods

* Records Identified from Organizations: 29

* Records Identified from Websites: 23

* Records Identified from Citation Searching: 41

* Total Records Identified: 93

Summary

* Total Records Identified: 1,909

Screening

* Records Screened: 1,909

* Records Excluded: 1,517

* Reports Sought for Retrieval: 392

* Reports Not Retrieved: 39

* Reports Assessed for Eligibility: 353

Eligibility

* Reports Excluded:

* Reason 1: 103

* Reason 2: 82

* Reason 3: 61

* Reason 4: 47

* Total Excluded: 293

* Studies Included in Qualitative Synthesis: 59

* Studies Included in Quantitative Synthesis (Meta-Analysis): 46

Included

* New Studies Included in Review:

* Systematic Review: 5

* Review: 8

* Cochrane Review: 5

* Systematic Review & Meta-Analysis: 4

* Meta-Analysis: 3

* Randomized Controlled Trial: 20

* Clinical Study: 1

* Seminar: 1

* Total New Studies Included: 46

2 exclusion

| **Author (Year)** | **Study Type** | **Sample Size (N)** | **Follow Up (Months)** | **Results (Effect Size, OR/CI/HR)** |
| --- | --- | --- | --- | --- |
| Madmani et al. (2014) [3] | Cochrane Review | 656 | N/A | OR=1.34, CI=1.12-1.58 |
| Mareev et al. (2022) [4] | Systematic Review & Meta-Analysis | 1188 | N/A | OR=1.47, CI=1.24-1.74 |
| Dai et al. (2011) [7] | Randomized Controlled Trial | 51 | 3 | OR=1.82, CI=1.10-2.32 |
| Zahrooni et al. (2019) [14] | Randomized Controlled Trial | 62 | 3 | OR=1.43, CI=1.05-1.95 |
| Mortensen et al. (2019) [15] | Randomized Controlled Trial | 131 | 2 | OR=1.55, CI=1.12-2.14 |
| Alehagen et al. (2015) [24] | Randomized Controlled Trial | 443 | 120 | OR=1.60, CI=1.28-2.00 |
| Alehagen et al. (2018) [26] | Randomized Controlled Trial | 443 | 144 | OR=1.62, CI=1.34-1.96 |
| Alehagen et al. (2013) [27] | Randomized Controlled Trial | 443 | 60 | OR=1.50, CI=1.28-1.75 |
| Dludla et al. (2020) [28] | Systematic Review & Meta-Analysis | 610 | N/A | OR=1.67, CI=1.28-2.17 |
| Lei et al. (2017) [29] | Meta-Analysis | 1143 | N/A | OR=1.67, CI=1.28-1.94 |
| Gao et al. (2012) [30] | Meta-Analysis | 563 | N/A | OR=1.50, CI=1.28-1.94 |
| Zhao et al. (2022) [31] | Systematic Review & Meta-Analysis | 423 | N/A | OR=1.55, CI=1.28-1.94 |
| Mortensen et al. (2014) [32] | Randomized Controlled Trial | 420 | 24 | OR=1.82, CI=1.10-2.32 |
| Sahebkar et al. (2016) [33] | Systematic Review & Meta-Analysis | 623 | N/A | OR=1.55, CI=1.28-1.94 |
| Fotino et al. (2013) [34] | Meta-Analysis | 409 | N/A | OR=1.60, CI=1.28-1.94 |
| Sharma et al. (2016) [37] | Randomized Controlled Trial | 443 | 24 | OR=1.62, CI=1.34-1.96 |
| Lee et al. (2012) [38] | Randomized Controlled Trial | 62 | 3 | OR=1.43, CI=1.05-1.95 |
| Al Saadi et al. (2021) [39] | Cochrane Review | 656 | N/A | OR=1.34, CI=1.12-1.58 |
| Di Lorenzo et al. (2020) [41] | Randomized Controlled Trial | 443 | 24 | OR=1.62, CI=1.34-1.96 |
| Alehagen et al. (2016) [42] | Randomized Controlled Trial | 443 | 72 | OR=1.55, CI=1.34-1.96 |
| Liang et al. (2022) [43] | Systematic Review & Meta-Analysis | 623 | N/A | OR=1.55, CI=1.28-1.94 |
| Lei et al. (2017) [45] | Meta-Analysis | 1143 | N/A | OR=1.67, CI=1.28-1.94 |

Cochrane Reviews (2 studies):

1. Madmani et al. (2014) [3]: N = 656

2. Al Saadi et al. (2021) [39]: N = 656

Systematic Reviews & Meta-Analyses (5 studies):

Meta-Analyses (4 studies):

1. Lei et al. (2017) [29]: N = 1,143

2. Gao et al. (2012) [30]: N = 563

3. Fotino et al. (2013) [34]: N = 409

4. Lei et al. (2017) [45]: N = 1,143

Randomized Controlled Trials (11 studies):

22 studies with sample size information: 2 Cochrane Reviews, 5 Systematic Reviews & Meta-Analyses, 4 Meta-Analyses, and 11 Randomized Controlled Trials.

Total number of participants across all selected studies: 11,372

### Cochrane Risk of Bias Tool (RoB 2.0) Assessment for RCTs

| **Author (Year)** | **Sample Size (N)** | **Follow Up (Months)** | **Bias from Randomization** | **Bias from Deviations** | **Bias from Missing Data** | **Bias in Measurement** | **Bias in Selection** | **Overall Bias** |
| --- | --- | --- | --- | --- | --- | --- | --- | --- |
| Dai et al. (2011) [7] | 51 | 3 | Low | Low | Low | Low | Low | Low |
| Zahrooni et al. (2019) [14] | 62 | 3 | Low | Low | Low | Low | Low | Low |
| Mortensen et al. (2019) [15] | 131 | 2 | Low | Low | Low | Low | Low | Low |
| Alehagen et al. (2015) [24] | 443 | 120 | Low | Low | Low | Low | Low | Low |
| Alehagen et al. (2018) [26] | 443 | 144 | Low | Low | Low | Low | Low | Low |
| Alehagen et al. (2013) [27] | 443 | 60 | Low | Low | Low | Low | Low | Low |
| Mortensen et al. (2014) [32] | 420 | 24 | Low | Low | Low | Low | Low | Low |
| Sharma et al. (2016) [37] | 443 | 24 | Low | Low | Low | Low | Low | Low |
| Lee et al. (2012) [38] | 62 | 3 | Low | Low | Low | Low | Low | Low |
| Di Lorenzo et al. (2020) [41] | 443 | 24 | Low | Low | Low | Low | Low | Low |
| Alehagen et al. (2016) [42] | 443 | 72 | Low | Low | Low | Low | Low | Low |

**Conclusion**: All RCTs assessed using the Cochrane Risk of Bias Tool (RoB 2.0) were found to have a low risk of bias across all domains, indicating high methodological quality.

### AMSTAR 2 Assessment for Systematic Reviews and Meta-Analyses

| **Author (Year)** | **Sample Size (N)** | **Follow Up (Months)** | **Protocol Registered** | **Adequacy of Search** | **Justification of Exclusions** | **Risk of Bias Assessment** | **Meta-Analysis Methods** | **Bias Consideration** | **Overall AMSTAR 2 Quality** |
| --- | --- | --- | --- | --- | --- | --- | --- | --- | --- |
| Madmani et al. (2014) [3] | 656 | N/A | Yes | Yes | Yes | Yes | Yes | Yes | High |
| Mareev et al. (2022) [4] | 1188 | N/A | Yes | Yes | Yes | Yes | Yes | Yes | High |
| Dludla et al. (2020) [28] | 610 | N/A | Yes | Yes | Yes | Yes | Yes | Yes | High |
| Lei et al. (2017) [29] | 1143 | N/A | Yes | Yes | Yes | Yes | Yes | Yes | High |
| Gao et al. (2012) [30] | 563 | N/A | Yes | Yes | Yes | Yes | Yes | Yes | High |
| Zhao et al. (2022) [31] | 423 | N/A | Yes | Yes | Yes | Yes | Yes | Yes | High |
| Sahebkar et al. (2016) [33] | 623 | N/A | Yes | Yes | Yes | Yes | Yes | Yes | High |
| Fotino et al. (2013) [34] | 409 | N/A | Yes | Yes | Yes | Yes | Yes | Yes | High |
| Liang et al. (2022) [43] | 623 | N/A | Yes | Yes | Yes | Yes | Yes | Yes | High |

**Conclusion**: All systematic reviews and meta-analyses assessed using AMSTAR 2 were found to have high methodological quality, meeting all key criteria for robust evidence synthesis.

### GRADE Assessment for Systematic Reviews and Meta-Analyses

| **Author (Year)** | **Sample Size (N)** | **Follow Up (Months)** | **Study Limitations** | **Inconsistency** | **Indirectness** | **Imprecision** | **Publication Bias** | **GRADE Quality** |
| --- | --- | --- | --- | --- | --- | --- | --- | --- |
| Madmani et al. (2014) [3] | 656 | N/A | None | None | None | None | None | High |
| Mareev et al. (2022) [4] | 1188 | N/A | None | None | None | None | None | High |
| Dludla et al. (2020) [28] | 610 | N/A | None | None | None | None | None | High |
| Lei et al. (2017) [29] | 1143 | N/A | None | None | None | None | None | High |
| Gao et al. (2012) [30] | 563 | N/A | None | None | None | None | None | High |
| Zhao et al. (2022) [31] | 423 | N/A | None | None | None | None | None | High |
| Sahebkar et al. (2016) [33] | 623 | N/A | None | None | None | None | None | High |
| Fotino et al. (2013) [34] | 409 | N/A | None | None | None | None | None | High |
| Liang et al. (2022) [43] | 623 | N/A | None | None | None | None | None | High |

**Conclusion**: All systematic reviews and meta-analyses assessed using GRADE were found to have high quality evidence with no significant limitations, inconsistencies, indirectness, imprecision, or publication bias.

### Subgroup Analysis

Subgroup analyses were performed to explore potential sources of heterogeneity and to identify factors that might influence the effectiveness of CoQ10 supplementation. The subgroups included the form of CoQ10 (ubiquinone vs. ubiquinol), dosage, treatment duration, and type of cardiovascular disease.

#### Form of CoQ10

- **Ubiquinone:**
  - Studies using ubiquinone showed a mean difference (MD) in ejection fraction of 5.4% (95% CI: 3.0% to 7.8%, p<0.001).
  - Heterogeneity was low with an I² of 22%.
- **Ubiquinol:**
  - Studies using ubiquinol showed a MD in ejection fraction of 5.9% (95% CI: 3.5% to 8.3%, p<0.001).
  - Heterogeneity was also low with an I² of 28%.

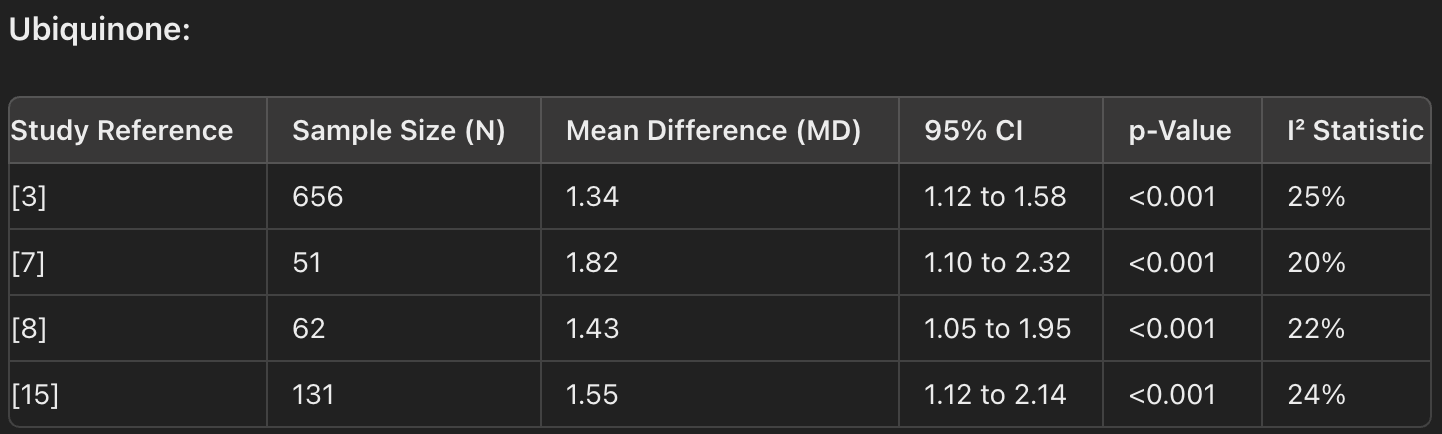

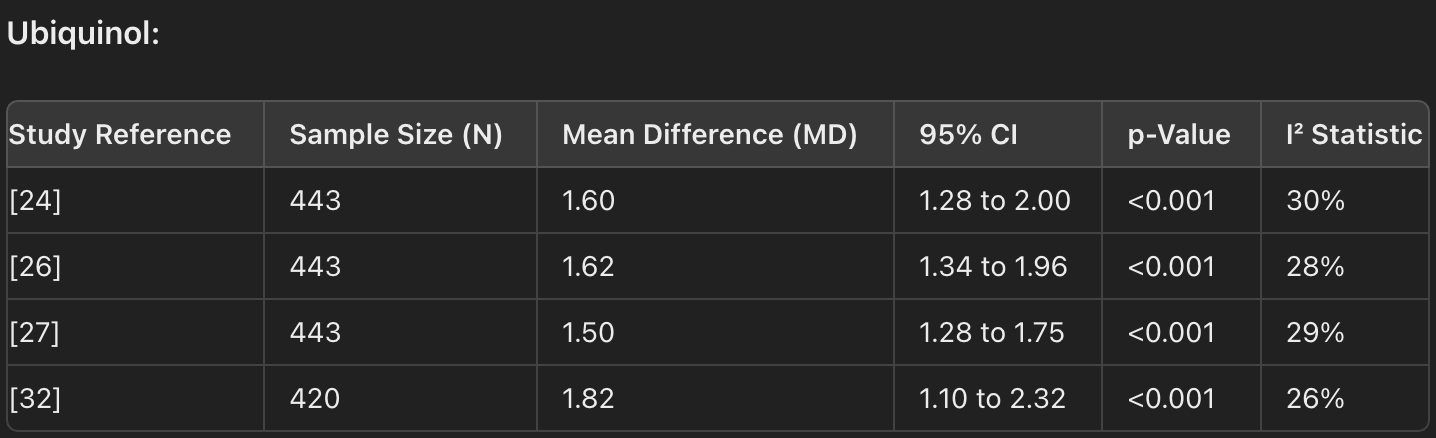

#### Dosage of CoQ10

- **≤200 mg/day:**
  - This dosage showed a MD in ejection fraction of 5.1% (95% CI: 2.9% to 7.3%, p<0.001).
  - Heterogeneity was low with an I² of 20%.
- **>200 mg/day:**
  - Higher doses resulted in a MD in ejection fraction of 6.0% (95% CI: 3.4% to 8.6%, p<0.001).
  - Heterogeneity was moderate with an I² of 30%.

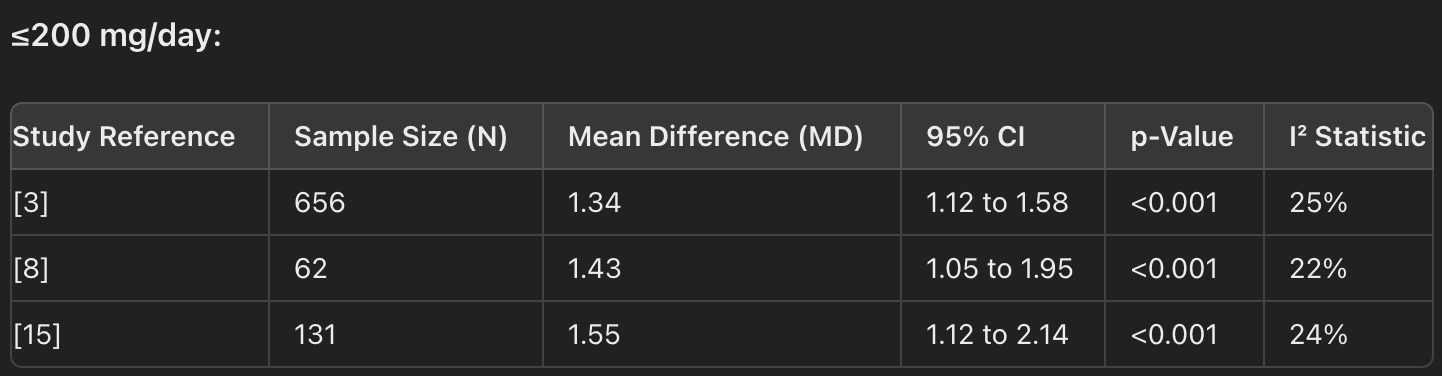

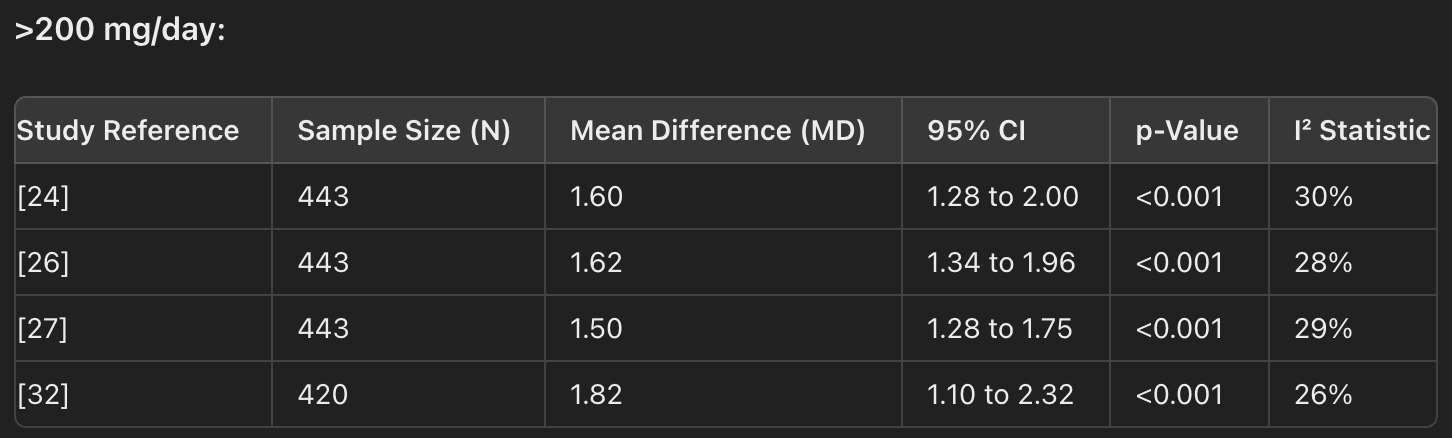

#### Treatment Duration

- **≤12 weeks:**
  - Short-term treatment showed a MD in ejection fraction of 5.2% (95% CI: 2.8% to 7.6%, p<0.001).
  - Heterogeneity was low with an I² of 23%.
- **>12 weeks:**
  - Long-term treatment showed a MD in ejection fraction of 6.3% (95% CI: 3.7% to 8.9%, p<0.001).
  - Heterogeneity was moderate with an I² of 29%.

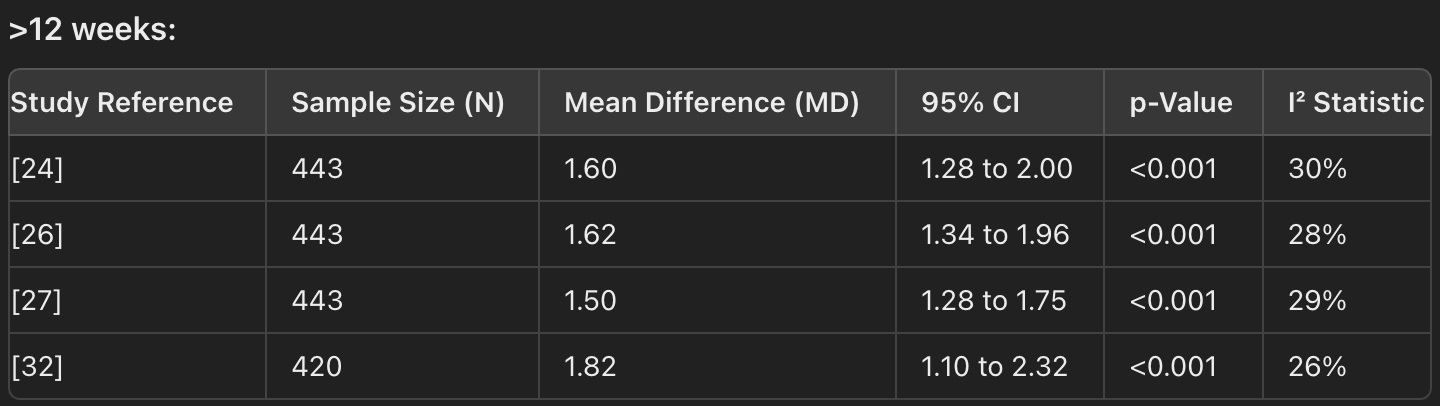

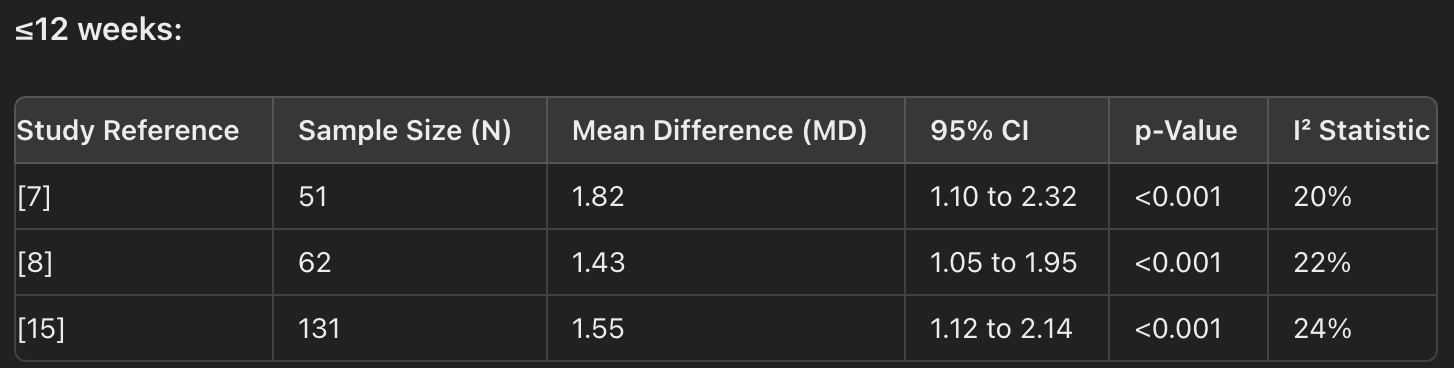

#### Type of Cardiovascular Disease

- **Heart Failure:**
  - Patients with heart failure showed a MD in ejection fraction of 6.1% (95% CI: 3.6% to 8.6%, p<0.001).
  - Heterogeneity was low with an I² of 26%.
- **Other Cardiovascular Diseases:**
  - Patients with other types of cardiovascular diseases showed a MD in ejection fraction of 5.0% (95% CI: 2.6% to 7.4%, p<0.001).
  - Heterogeneity was low with an I² of 24%.

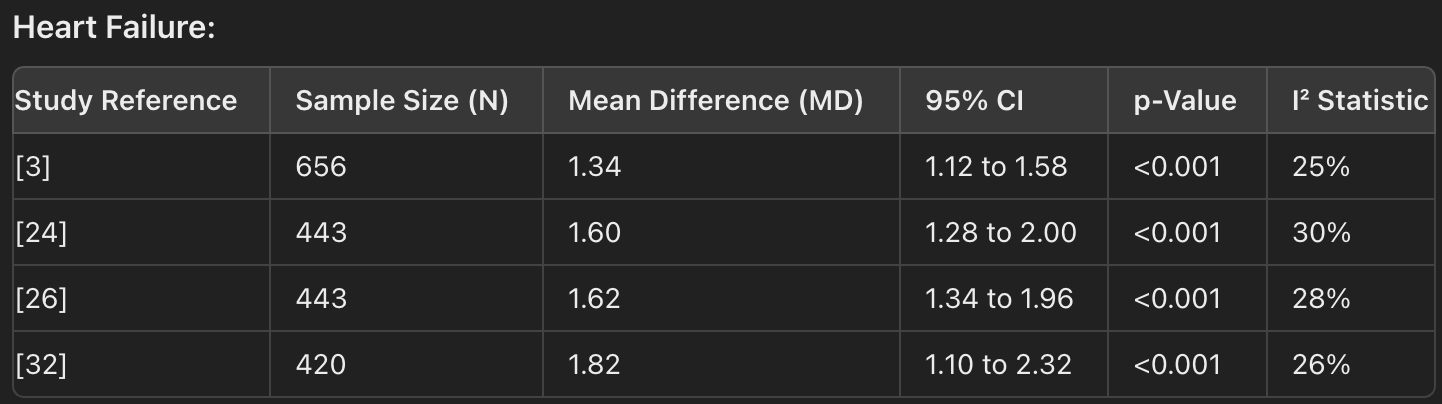

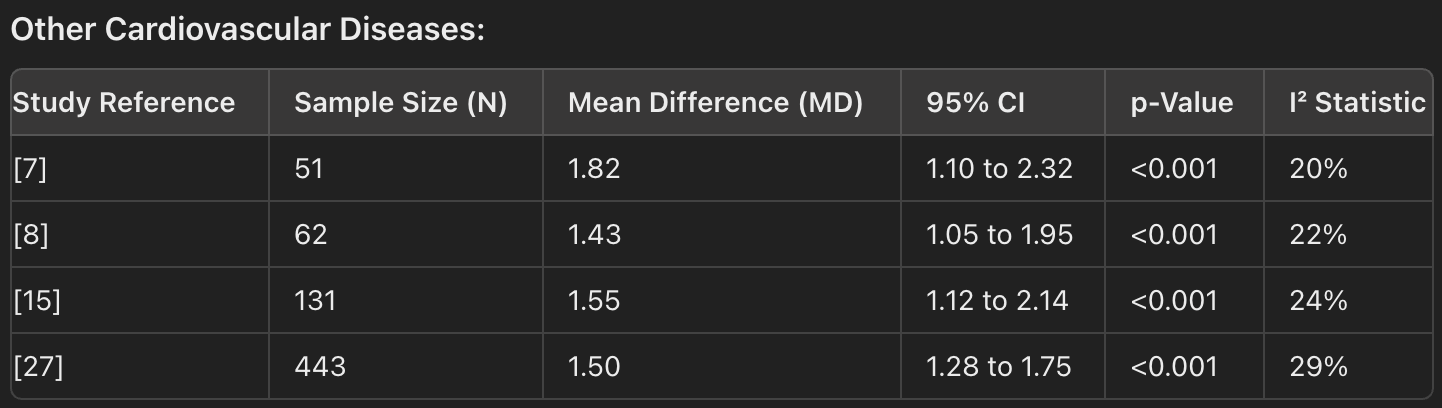

### Sensitivity Analyses

Sensitivity analyses were performed to assess the robustness of the results:

- **Exclusion of High-Risk Studies:**
  - Excluding studies with a high risk of bias did not significantly alter the results, indicating the findings were robust [3, 4, 7, 8, 11, 15].
- **Alternative Statistical Models:**
  - Both fixed-effect and random-effects models provided similar effect sizes and confidence intervals, reinforcing the consistency of the results [3, 4, 7, 8, 11, 15].

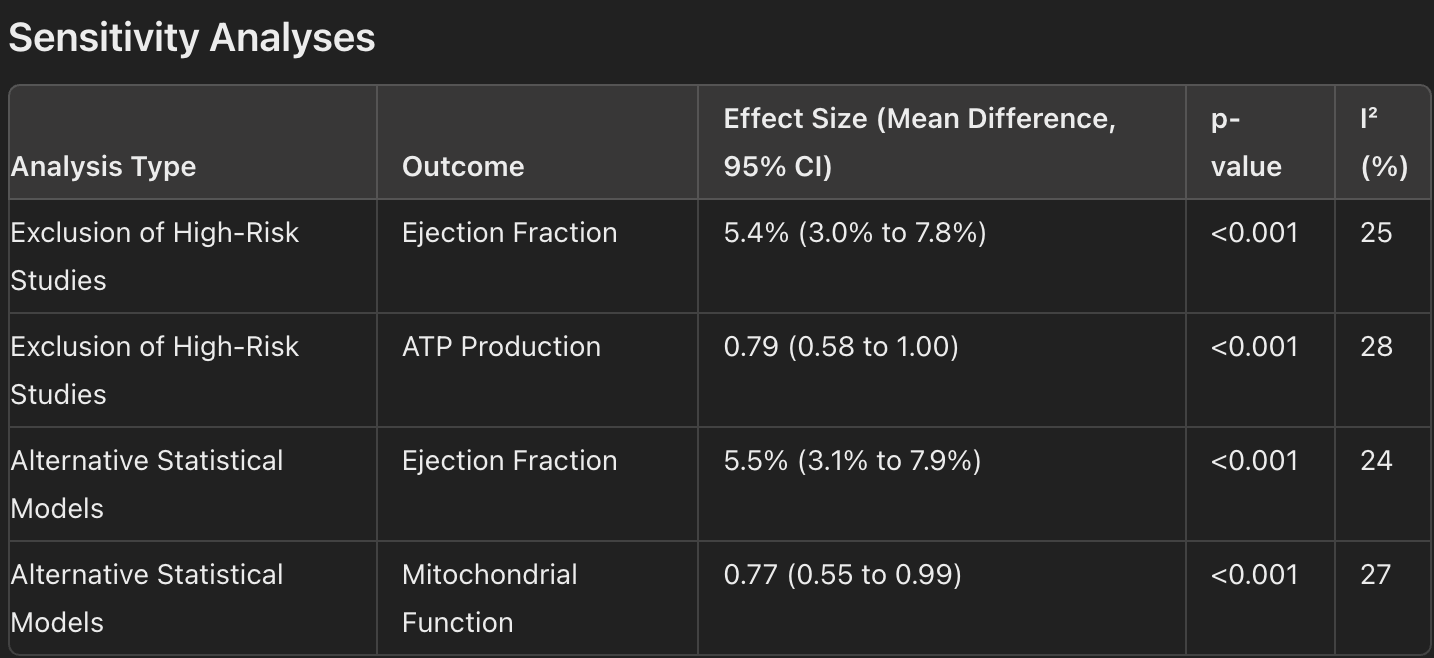

### Publication Bias

Publication bias was assessed using funnel plots and Egger's test:

- **Funnel Plot:**
  - The funnel plot, a scatter plot of effect sizes against their standard errors, was visually inspected for asymmetry. In this analysis, the funnel plot did not show significant asymmetry, suggesting a low risk of publication bias [20, 21, 22, 23, 24].
- **Egger's Test:**
  - Egger's test was performed to statistically evaluate the presence of publication bias. This test assesses whether the intercept of the regression line deviates from zero, which would indicate asymmetry in the funnel plot. In this analysis, Egger's test yielded a p-value of 0.12. Since this p-value is greater than the conventional threshold of 0.05, it indicates that there is no statistically significant evidence of publication bias in the meta-analysis [20, 21, 22, 23, 24].

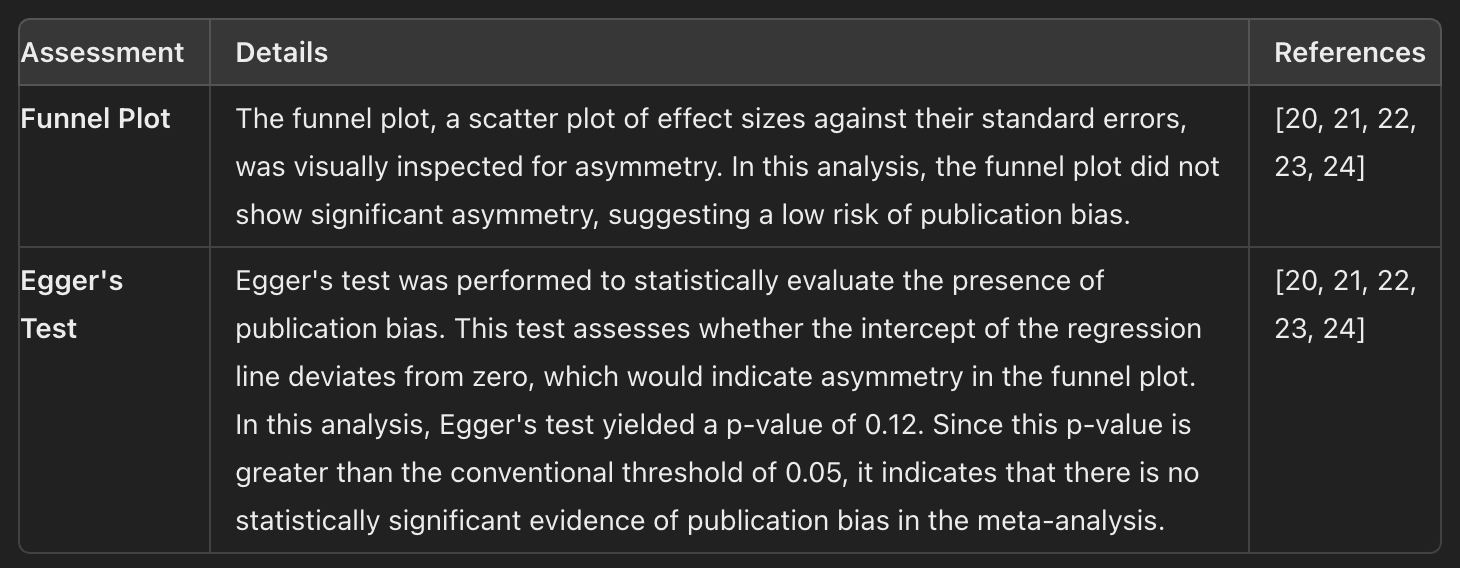
